# Population-level effectiveness of pre-exposure prophylaxis for HIV prevention among men who have sex with men in Montréal: a modelling study of surveillance and survey data

**DOI:** 10.1101/2023.05.31.23290795

**Authors:** Carla M Doyle, Rachael M Milwid, Joseph Cox, Yiqing Xia, Gilles Lambert, Cécile Tremblay, Joanne Otis, Marie-Claude Boily, Jean-Guy Baril, Réjean Thomas, Alexandre Dumont Blais, Benoit Trottier, Daniel Grace, David M. Moore, Sharmistha Mishra, Mathieu Maheu-Giroux

## Abstract

**Background:** HIV pre-exposure prophylaxis (PrEP) has been recommended and partly subsidized in Québec since 2013. We aimed to evaluate the population-level impact of PrEP on HIV transmission among men who have sex with men (MSM) in Montréal over 2013-2021.

**Methods:** We used an agent-based mathematical model of sexual HIV transmission to estimate the fraction of HIV acquisitions averted by PrEP compared to a counterfactual scenario without PrEP. The model was calibrated to local MSM survey and cohort data and accounted for COVID-19 pandemic impacts on sexual activity, prevention, and care. To assess potential optimization strategies, we modelled hypothetical scenarios prioritizing PrEP to MSM with high sexual activity or aged ≤45 years, increasing coverage to levels achieved in Vancouver (where PrEP is free-of-charge), and improving retention.

**Results:** Over 2013-2021, the estimated annual HIV incidence decreased from 0.4 (90% credible interval [CrI]: 0.3-0.6) to 0.2 (90%CrI: 0.1-0.2) per 100 person-years. PrEP coverage in HIV-negative MSM remained low until 2015 (<1%). Afterward, coverage increased to a maximum of 10% (15% of those eligible for PrEP) and the cumulative fraction of HIV acquisitions averted over 2015-2021 was 20% (90%CrI: 11%-30%). The hypothetical scenarios modelled showed that PrEP could have averted up to 63% (90%CrI: 54%-70%) of acquisitions if coverage reached 10% in 2015 and 30% in 2019, like in Vancouver.

**Interpretation:** PrEP reduced population-level HIV transmission among Montréal MSM. However, our study suggests missed prevention opportunities and provides support for public policies that provide PrEP free-of-cost to MSM at high risk of HIV acquisition.

## Introduction

After over 20 years under study^1-3^ and ten years of availability^4^, oral pre-exposure prophylaxis (PrEP) has proven highly efficacious for preventing HIV acquisition across transmission routes. Rigorous randomized controlled clinical and pragmatic trials and other observational data speak directly to its prevention benefits among men who have sex with men (MSM) when taken daily or on-demand^5-10^. The IPERGAY trial in Canada (Québec) and France showed an 86% (95% confidence interval [CI]: 40%-98%) efficacy in preventing HIV acquisition among MSM assigned to on-demand oral PrEP compared to placebo^6^. Furthermore, its open-label extension study suggested that effectiveness could increase to 97% (95%CI: 81%-100%) among fully adherent on-demand PrEP users^8^. However, limited research as examined PrEP’s real-world impact on HIV dynamics over years of implementation, and all existing studies were empirical studies relying on observed HIV diagnoses^11, 12^, which can be affected by testing efforts.

Individual-based trials are important to demonstrate individual-level effectiveness but cannot provide the effect size estimates of public health relevance: the population-level relative decline in HIV incidence. Beyond the direct prevention benefits to PrEP users, the population-level effects include the indirect gains accrued by individuals not taking PrEP, who are at reduced risk of HIV acquisition as PrEP interrupts transmission chains and diminishes the number of their contacts able to transmit HIV. However, estimating this population-level effect empirically can be methodologically challenging, as the ongoing dynamics of HIV transmission and the use of other prevention tools can make it difficult to define suitable control groups. In Québec, concomitant 2013-2015 changes in antiretroviral treatment (ART) guidelines to immediate initiation and strengthened “undetectable=untransmittable” messaging especially complicate this. Under these circumstances, mathematical modelling can be advantageous^13^.

Québec implemented interim guidelines for tenofovir disoproxil fumarate/emtricitabine (TDF-FTC) as HIV PrEP in 2013^14^, the only province/territory to do so ahead of Health Canada’s licencing in 2016 and the development of national PrEP guidelines in 2017^15^. These guidelines recommended PrEP for MSM that had condomless anal sex in the past six months and met one of the following criteria related to sexual behaviour: 1) two or more sex partners in the past six months, 2) history of repeated post-exposure prophylaxis (PEP) use, 3) history of syphilis or an anal bacterial sexually transmitted infection, 4) sex with a partner living with HIV whose risk of transmission is considered high, or 5) psychoactive substance use during sex^14, 16, 17^. At the same time, Québec’s drug insurance program included TDF-FTC as PrEP in its formulary, reducing its cost to a monthly co-payment of up to CAD$96.74^18^. However, nearly a decade later, PrEP’s impact on the local HIV epidemic has not yet been evaluated, partly due to the aforementioned challenges. Such evaluations are essential for understanding PrEP’s role in eliminating HIV as a public health threat and improving its delivery.

With over two million people, Montréal is Québec’s largest city^19^ and the epicentre of its HIV epidemic^20^. It was the first city in Canada to join the Fast-Track Cities initiative, aiming to eliminate HIV^21^, and has well-established HIV surveillance and population-based data for monitoring the HIV response. Leveraging these data, we evaluated the population-level effectiveness of PrEP on HIV transmission among MSM in Montréal over 2013-2021 and investigated if and how this intervention could have been optimized. Our analysis employed a mathematical model to disentangle the unique contribution of PrEP from other interventions and simulate an appropriate counterfactual scenario. PrEP is pillar of HIV elimination efforts^22^ and preferred prevention method for many Canadian MSM^23^. Understanding the impact of PrEP can guide decision-makers in accelerating the city’s progress toward zero new HIV acquisitions.

## Methods

### Model overview

We used an existing calibrated agent-based model of sexual HIV transmission among Montréal MSM, described elsewhere^24^. Briefly, it is a stochastic, mechanistic model including several modules to simulate demographics, partnership formation and dissolution (i.e., casual and regular sex partnerships, and mixing by age, serostatus, and preferred insertive/receptive role during anal sex), use of HIV prevention tools (i.e., condoms, PEP, PrEP, and viral suppression), the HIV treatment and care cascade (i.e., HIV testing and ART), HIV transmission, and disease progression^24^. Initialized in 1975, the model tracks a population of 10,000 MSM aged 15+ years that increases over time, reflecting Montréal’s demographics. Men are categorized by age (15-24, 25-34, 35-44, 45-54, 55+ years) and sexual activity level (low, medium, and high, with 0-5, 6-10, and 11+ sexual partners per year, respectively), and exit the model due to death from natural or HIV/AIDS-related causes. The model is implemented in R (v.4.1.0) with a C++ back-end using the Rcpp library^25-27^, and simulated with a two-week time step.

We calibrated the model to the following sexual behaviour, epidemic, and intervention outcomes: the distribution of the number of anal sex partners in the past six months, prevalence and duration of regular partnerships, CD4 cell count at diagnosis by year (2013-2017), HIV prevalence by age (18-29, 30-49, 50+ years) and year (2005, 2008, 2017-2019), the prevalence of lifetime PrEP use by year (2017-2019), PrEP coverage (defined as the proportion currently using PrEP among those not living with HIV) by year (2017-2019), the proportion of people living with HIV (PLHIV) diagnosed by year (2005, 2008, 2017), and ART coverage among PLHIV by year (2005, 2017-2018). Using an Approximate Bayesian Computational Sequential Monte Carlo fitting method^28, 29^, we obtained 100 calibrated parameter sets of the 54 parameters governing transmission and can reproduce the observed epidemic dynamics.

### PrEP-related data sources

We used data from two sources to parameterize and calibrate the PrEP module (Table 1). The *Engage Cohort*^30^, a population-based study of sexually active MSM aged 16+ years in Montréal, Toronto, and Vancouver, provided data from 2017-2021. In Montréal, a closed cohort of men that had sex with another man in the past six months were recruited over 2017-2018 (N=1,179) by respondent-driven sampling (RDS) and visits occurred annually until 2021. The *l’Actuel PrEP Cohort*^31^, an ongoing clinical cohort at Montréal’s *Clinique l’Actuel*, provided data on individuals consulting for and prescribed PrEP from 2013-2019 (N=2,746, 98% of which are MSM).

**Table 1:**
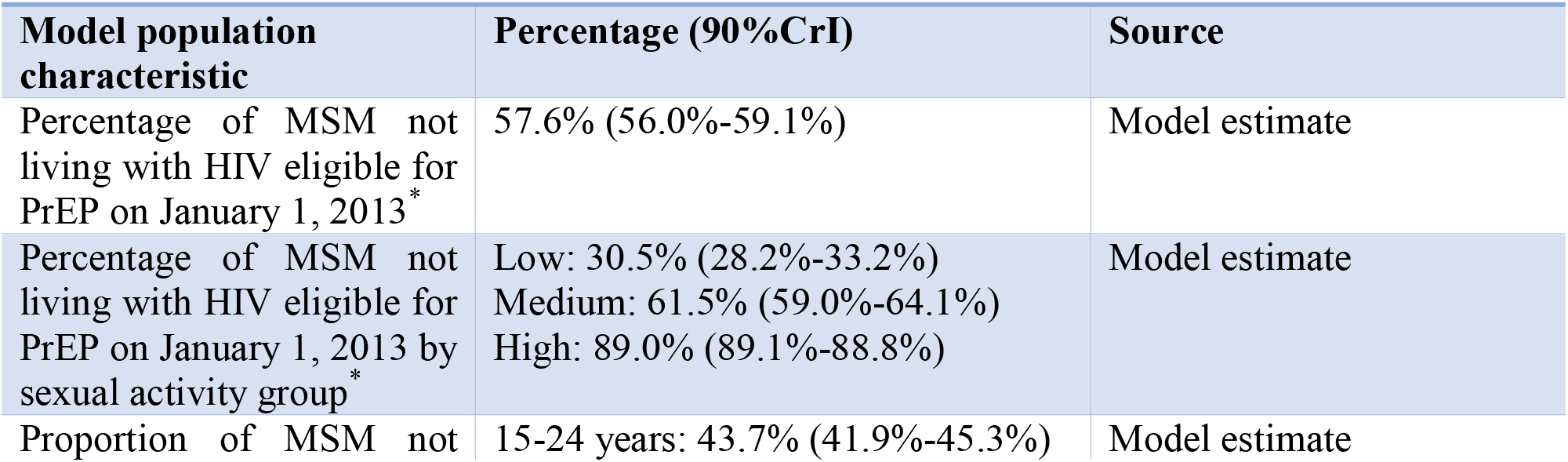
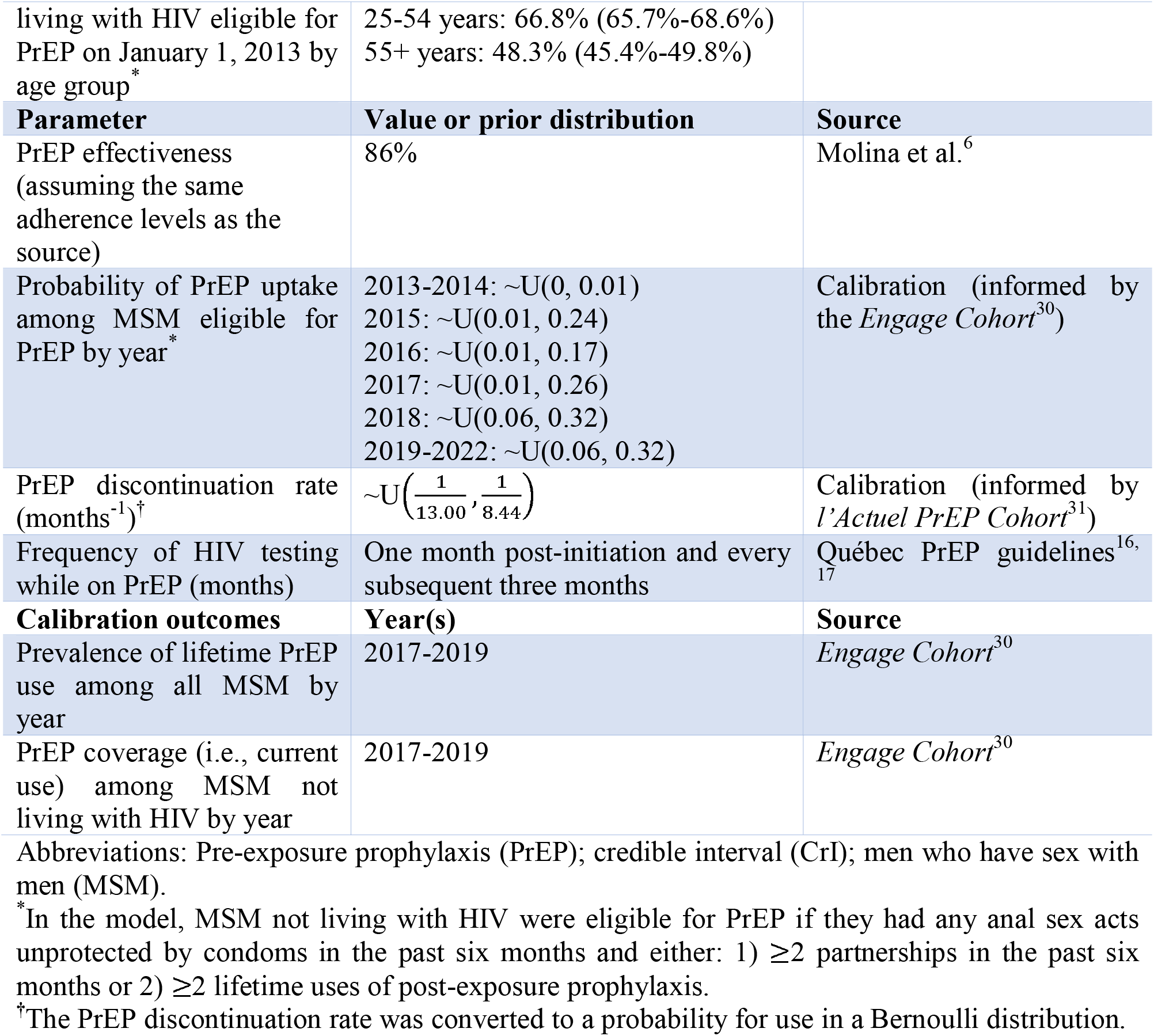
Summary of key PrEP-related model population characteristics, parameters, and calibration outcomes.

### PrEP parameterization

The PrEP module required effectiveness, uptake, and discontinuation parameters (Table 1). To parameterize effectiveness, we reviewed published literature and selected the IPERGAY trial’s intention-to-treat estimate of 86%, considering its relevance to Québec and accounting for imperfect PrEP adherence. We estimated the remaining parameters by calibrating to *Engage* data on lifetime PrEP use among all MSM and PrEP coverage among those not living with HIV.

We calibrated annual probabilities of PrEP uptake among PrEP-eligible men with prior distributions informed by *Engage. Engage* captured self-reported information on the year of first PrEP use among participants who reported ever using PrEP at baseline and using PrEP in the past six months during follow-up visits. We estimated the proportion of PrEP-eligible men that first took PrEP each year, accounting for the complex survey design using RDS-II sampling weights^32^ and for loss-to-follow-up using inverse probability of censoring weights (Figures S1-S2). To obtain estimates before 2017, we assumed the number eligible at baseline was constant. For calibration, we considered the 95%CI bounds and made additional assumptions. First, since no participant reported starting PrEP in 2013 and only seven reported starting in 2014, we assumed uptake was equivalent in those years and informed the prior by the 95%CI of the 2014 estimate. Second, there was increased uncertainty regarding attrition and measurement of PrEP initiation at the last study visit, possibly impacting the estimate for 2019. Since the 95%CI of the 2018 estimate was wide and included plausible values for both years, we used this to inform the prior distribution of the uptake probability in 2019. Finally, we incorporated additional uncertainty in all uptake probability prior distributions to allow for any residual bias due to attrition (Table 1).

Clinical data from l’*Actuel* informed the PrEP discontinuation rate prior distribution. We defined PrEP discontinuation based on three criteria: 1) reported stopping at a follow-up visit, 2) absence of reported stopping but undergoing another PrEP consultation, or 3) >180 days between visits. The discontinuation date was determined by the available data, as follows: 1) the reported stop date, 2) the date of the visit where stopping was reported, or 3) 3-6 months (randomly chosen from a uniform distribution) after the last visit before stopping. PrEP retention (i.e., duration of continuous use) was calculated as the time between initiation and discontinuation. The prior bounds of the discontinuation rate were obtained by inverting the age-standardized interquartile range of individual retention (Table 1).

### Modelling of PrEP initiation and discontinuation

Starting from 2013, the model simulated oral PrEP use. Matching the Québec PrEP guidelines, men susceptible to HIV acquisition were eligible for PrEP if they had any anal sex acts unprotected by condoms in the past six months and either: 1) ≥2 partnerships in the past six months or 2) ≥2 uses of lifetime PEP uses^16, 17^.

At each time step, a Bernoulli distribution using the calibrated uptake probabilities determined if each eligible man would initiate PrEP. Those selected for PrEP underwent HIV testing and started PrEP if the result was negative. HIV testing while on PrEP occurred after the first month of use and every subsequent three months. Those who tested positive for HIV immediately discontinued PrEP and were linked to care.

Finally, the calibrated discontinuation rate was converted to a probability and used in a Bernoulli distribution to determine which PrEP users would discontinue at each time step.

### Impacts of COVID-19 pandemic disruptions on sexual behaviours and PrEP

The COVID-19 pandemic’s impact was incorporated into the model starting from March 2020. Sexual activity, prevention, and treatment changes were informed by *Engage* data. PrEP use changes were also informed by *l’Actuel*. From March to June 2020, partner change rates decreased, with a 0.5 and 0.2 absolute reduction in the mean number of annual partners for those living and not living with HIV, respectively, in the low-medium sexual activity group and 5.0 and 10.4 for those living and not living with HIV, respectively, in the high sexual activity group. Due to service disruptions, reductions in the probabilities of testing annually (by 51% and 21% in the low-medium and high sexual activity groups, respectively), PEP initiation (by 43%), PrEP initiation (by 35%), and PrEP retention (discontinuation probability increased by 153%) remained until July 2021. After this time, we assumed a return to pre-pandemic levels.

### Model outputs and impact evaluation measures

We tracked characteristics of PrEP use and coverage (percentage of susceptible individuals taking PrEP) and HIV acquisitions over time. We then calculated the annual and cumulative numbers of HIV acquisitions. Finally, we estimated the annual incidence risk by the number of HIV acquisitions each year divided by the number susceptible to HIV acquisition at year-start.

The HIV epidemic was simulated ten times for each of the 100 calibrated parameter sets and the outputs were summarized by the mean. We performed this process under two scenarios: the provincial PrEP intervention scenario and the counterfactual scenario without PrEP. We measured PrEP’s population-level impact by the fraction of HIV acquisitions averted by PrEP over the total number of acquisitions in the counterfactual scenario without PrEP.

### Sensitivity analyses

In sensitivity analyses, we relaxed the eligibility criteria since MSM may have received PrEP despite not meeting all the provincial criteria^33^. We considered two scenarios, each applying only one eligibility criterion at a time: 1) condomless anal sex in the past six months, and 2) ≥2 partnerships in the past six months. The PrEP uptake rates were kept as calibrated.

Additionally, we assessed the robustness of our results to the assumed PrEP effectiveness, increasing it to 96%, as data indicated high adherence among continuous users (supplementary materials).

### Alternative PrEP intervention scenarios

We simulated alternative, hypothetical intervention scenarios to explore how PrEP’s impact could have been optimized (Table 2). These involved assessing the potential impact of PrEP prioritization (either to men in the high sexual activity group or those aged 45 years), increased coverage levels (up to a maximum of 30% by 2019, approximating the coverage reached in Vancouver, where PrEP is free for eligible individuals), and increased retention (reducing the discontinuation probability by 25% and 50%).

**Table 2.**
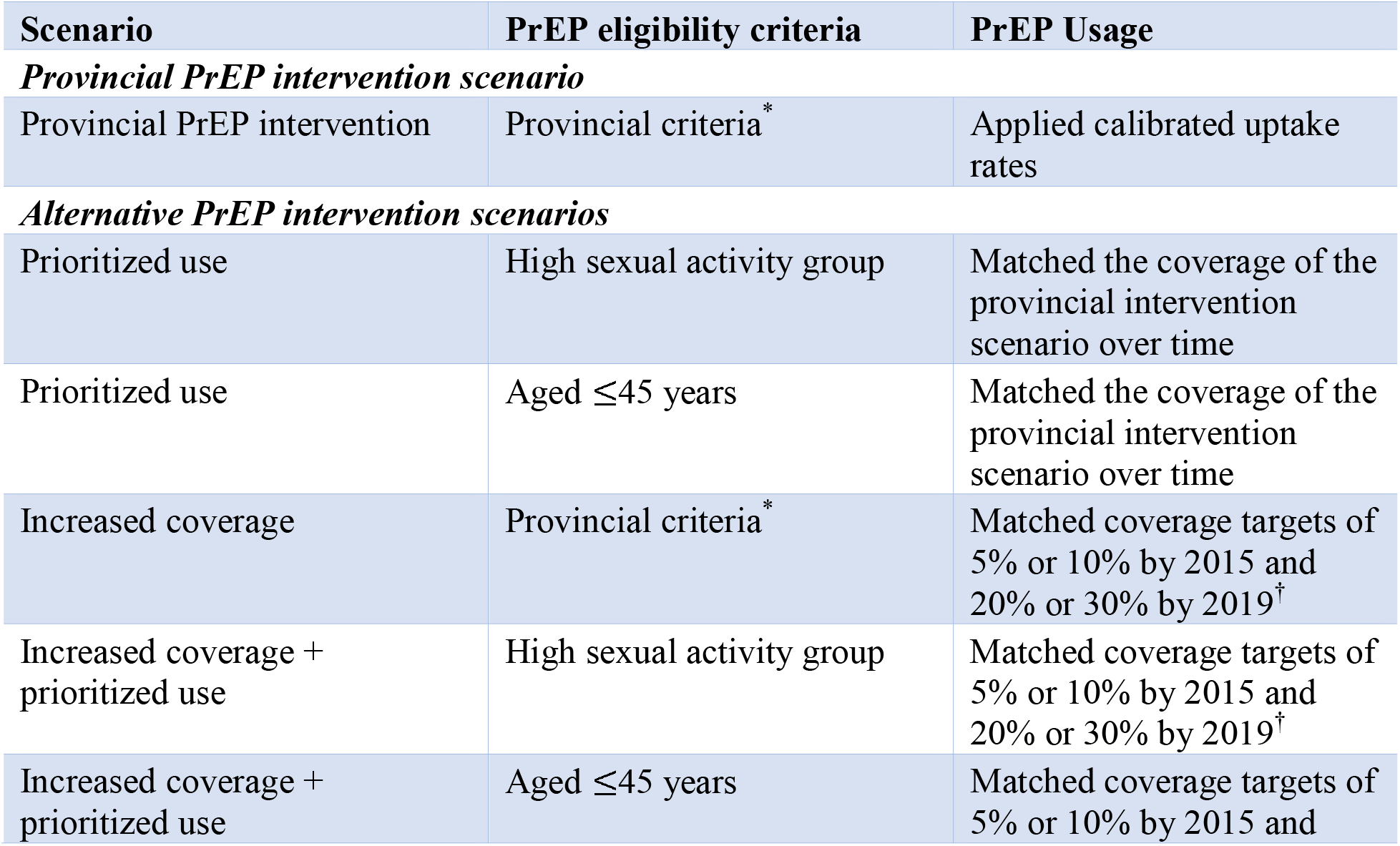
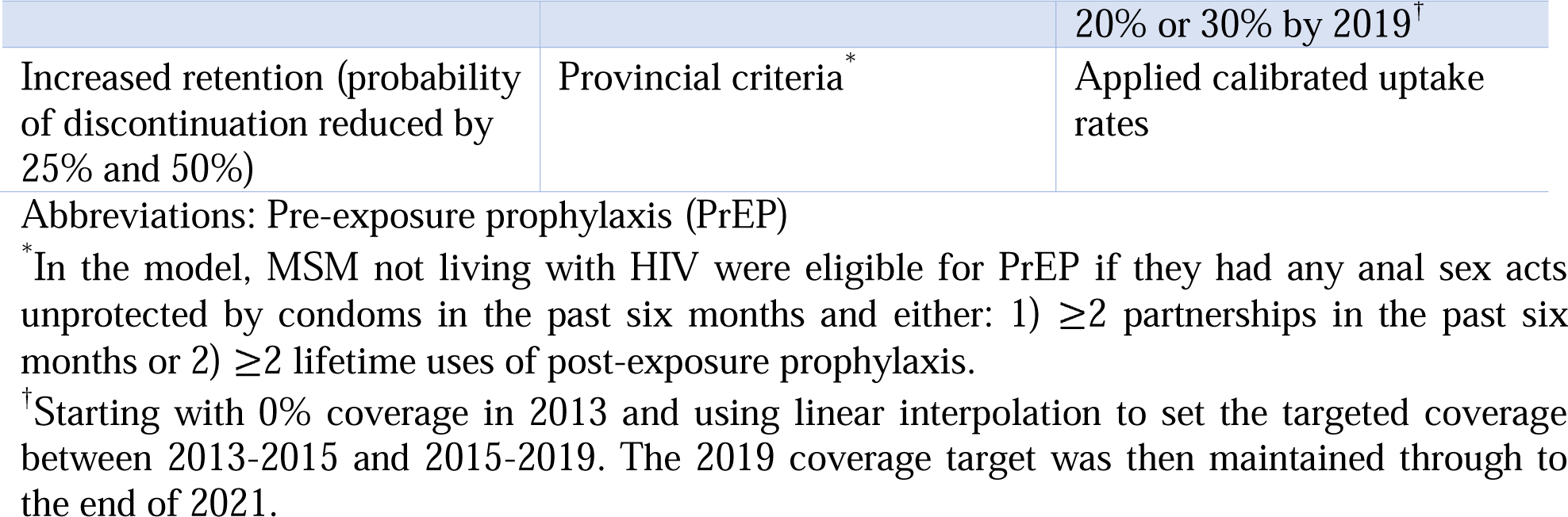
Pre-exposure prophylaxis (PrEP) intervention scenarios modeled among men who have sex with men in Montréal over 2013-2021.

### Ethics

The *McGill University Research Ethics Board* and the *Research Institute of the McGill University Health Centre* approved this study.

## Results

### PrEP coverage

According to *Engage* data, PrEP uptake was low after Québec’s interim guidelines were published in 2013 and gradually started increasing in 2015, and reaching 5% (95%CI: 3%-7%) coverage among MSM not living with HIV by 2018 (Figure 1). In 2020, *Engage* data indicated 10% (95%CI: 6%-16%) of Montréal MSM not living with HIV were currently on PrEP. Our model reflected these trends well, matching the estimated PrEP coverage in MSM not living with HIV at 4% (90% credible interval [CrI]: 2%-6%) in 2018 and 10% (90%CrI: 8%-12%) in 2020 (Figure 1). Among the PrEP-eligible population, which accounted for 62% (90%CrI: 61%-64%) of MSM not living with HIV (Figure S5), current use reached 16% (90%CrI: 13%-19%) in 2020 (Figure S6). During the initial waves of the COVID-19 pandemic, there was a significant decline in PrEP coverage due to reduced initiation and increased discontinuation (by model design), but coverage rebounded in mid-2021 (Figure 1). Throughout the study period, PrEP usage varied across different age and sexual activity groups (Figure 1).

**Figure 1.**
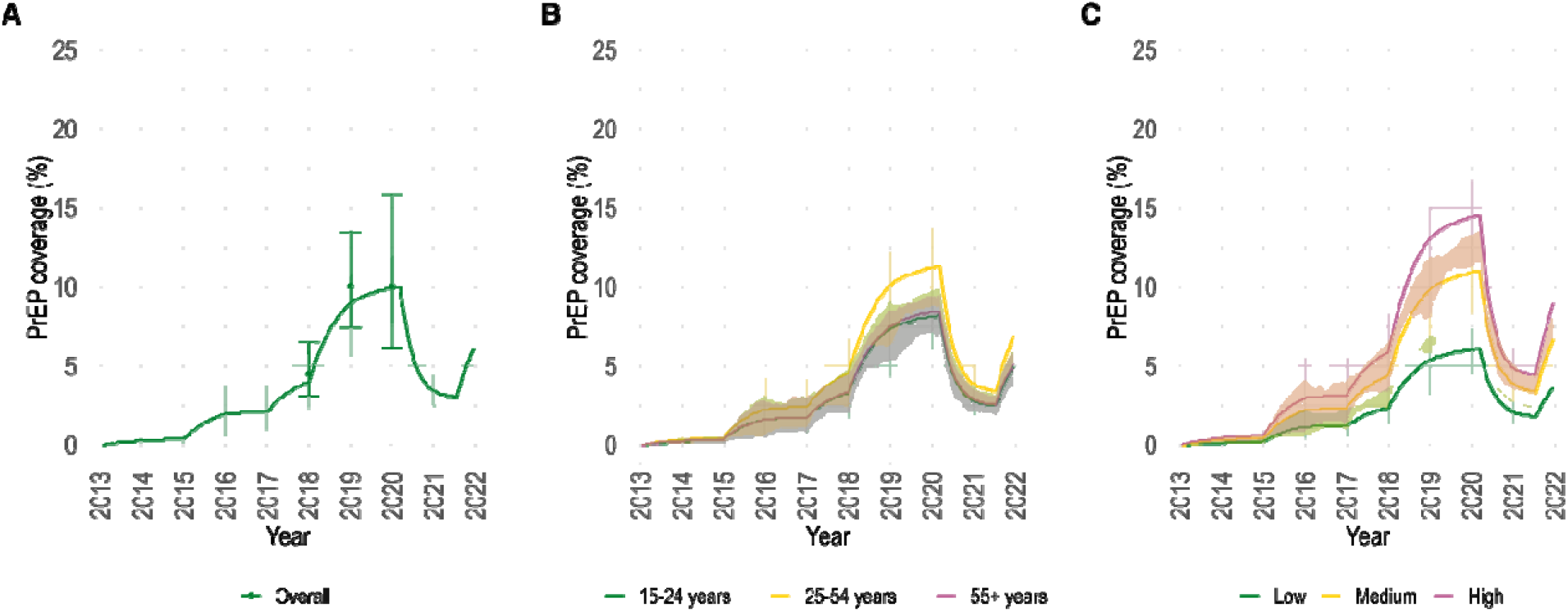
Pre-exposure prophylaxis (PrEP) coverage among men who have sex with men (MSM) not living with HIV in Montréal. Estimated PrEP coverage over 2013-2021 amon MSM not living with HIV in Montréal: overall (panel A) and stratified by age (panel B) and sexual activity group (panel C). The coloured lines and bands show the model posterior mean and 90% credible intervals, respectively. The three points and bars in panel A display the estimated PrEP coverage and 95% confidence intervals calculated from *Engage* and adjusted b RDS-II and inverse probability of censoring weights.

### Annual HIV incidence

The modelled annual HIV incidence was 0.4 per 100 person-years (90%CrI: 0.3-0.6) in 2013 and decreased to 0.2 per 100 person-years (90%CrI: 0.1-0.2) in 2021 (Figure 2). Prior to PrEP scale-up, the estimated annual incidence differed markedly by age and sexual activity levels (Figure 2). In 2013, the oldest age group (55+) had the lowest estimated annual incidence (0.2 per 100 person-years [90%CrI: 0.1-0.3]) compared to the 15-24-year-olds (0.3 per 100 person-years [90%CrI: 0.2-0.5]) and the 25-54-year-olds (0.5 per 100 person-years [90%CrI: 0.3-0.7]). By the end of 2020, incidence reached 0.1 per 100 person-years in the 15-24 (90%CrI: 0.1-0.2) and 55+ (90%CrI: 0-0.1) age groups, and 0.2 per 100 person-years (90%CrI: 0.1-0.3) in those aged 25-54. Across sexual activity levels, those with more sexual partners had a higher incidence. Over time, the most pronounced incidence reductions were exhibited by the highest sexual activity group, decreasing from 0.8 per 100 person-years (90%CrI: 0.6-1.1) in 2013 to 0.3 per 100 person-years (90%CrI: 0.2-0.5) in 2021. Considering PrEP use status over 2016-2021, incidence was higher among MSM eligible but not taking PrEP and even higher among those who discontinued PrEP (Figure 3).

**Figure 2.**
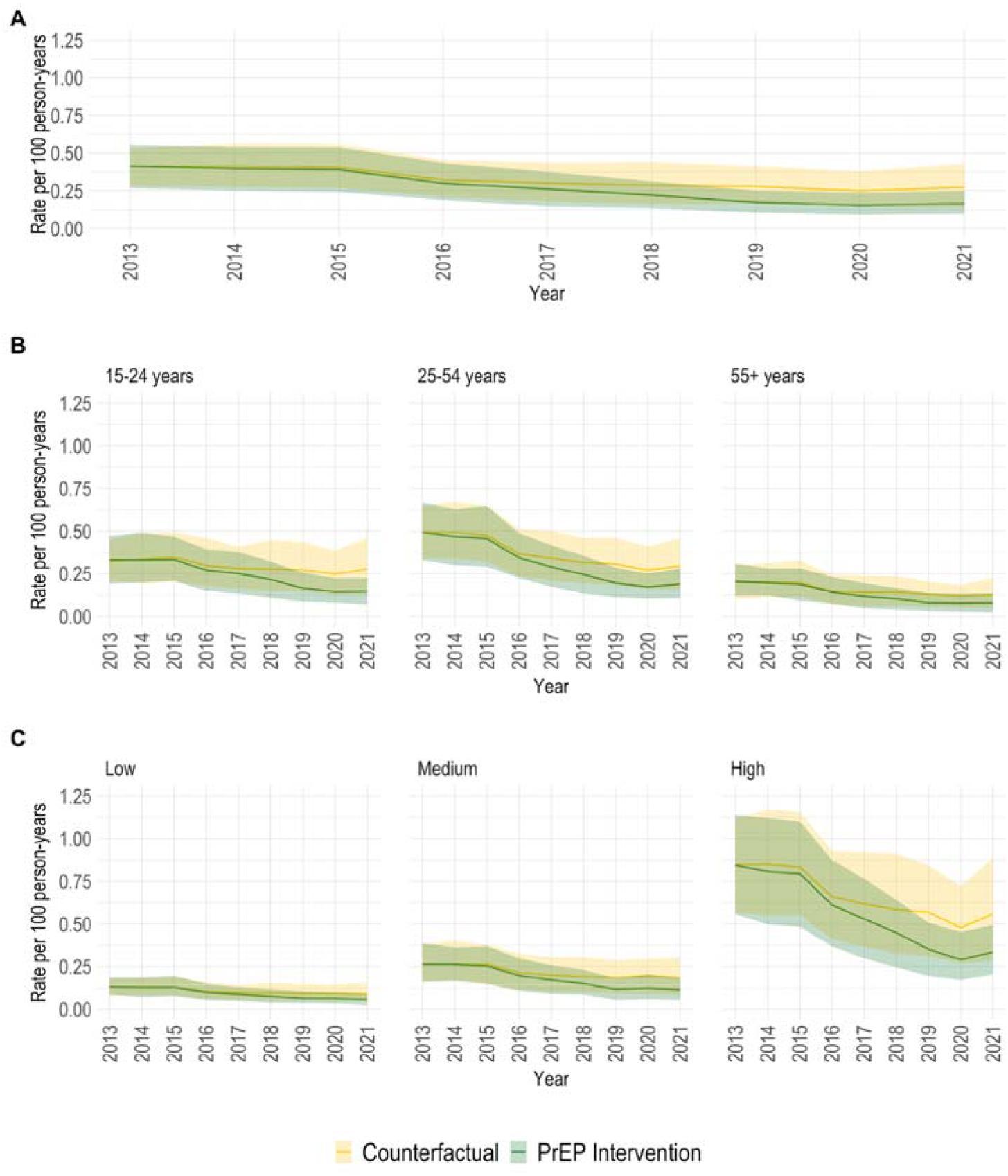
Annual HIV incidence in the provincial pre-exposure prophylaxis (PrEP) intervention and counterfactual scenarios. Estimated HIV incidence rates over 2013-2021 among men who have sex with men (MSM) in Montréal under the provincial pre-exposure prophylaxis (PrEP) intervention and counterfactual scenarios. The rates are presented overall (panel A) and stratified by age (panel B) and sexual activity group (panel C). The coloured lines and bands show the posterior mean and 95% credible intervals, respectively.

**Figure 3.**
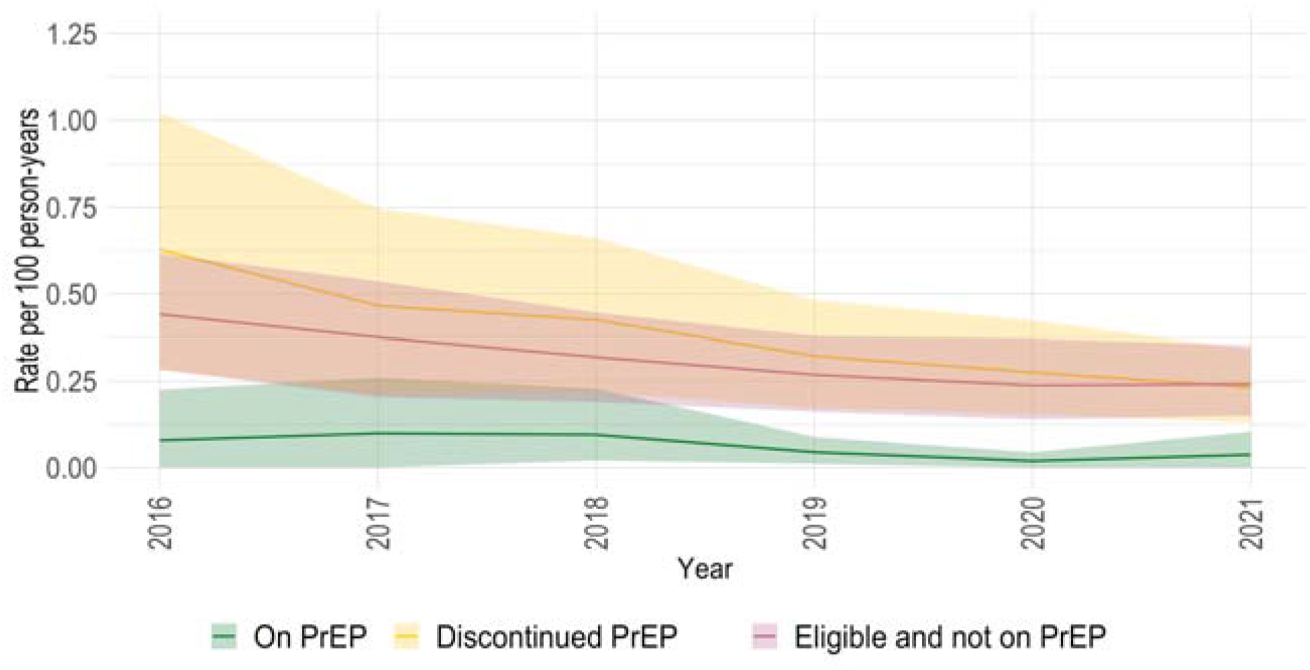
Annual HIV incidence by pre-exposure (PrEP) use status in the provincial PrEP intervention scenario. Estimated HIV incidence rates under the provincial pre-exposure prophylaxis (PrEP) intervention scenario over 2013-2021 among men who have sex with men (MSM) in Montréal, stratified by PrEP use status. The coloured lines and bands show the posterior mean and 95% credible intervals, respectively.

### Impact evaluation

Over the study period, the annual fractions of HIV acquisitions averted by PrEP increased (Figure 4). In the early years of PrEP availability, when coverage was lowest, it had a limited impact on averting HIV acquisitions. However, starting in 2017, PrEP began to have greater impacts. In 2021, PrEP averted an estimated 38% (90%CrI: 20%-53%) of HIV acquisitions. Given the low coverage before 2015, we focused the cumulative evaluation from 2015 onward and estimated that PrEP averted 20% (90%CrI: 11%-30%) of HIV acquisitions from 2015 to 2022 (Figure 4).

**Figure 4.**
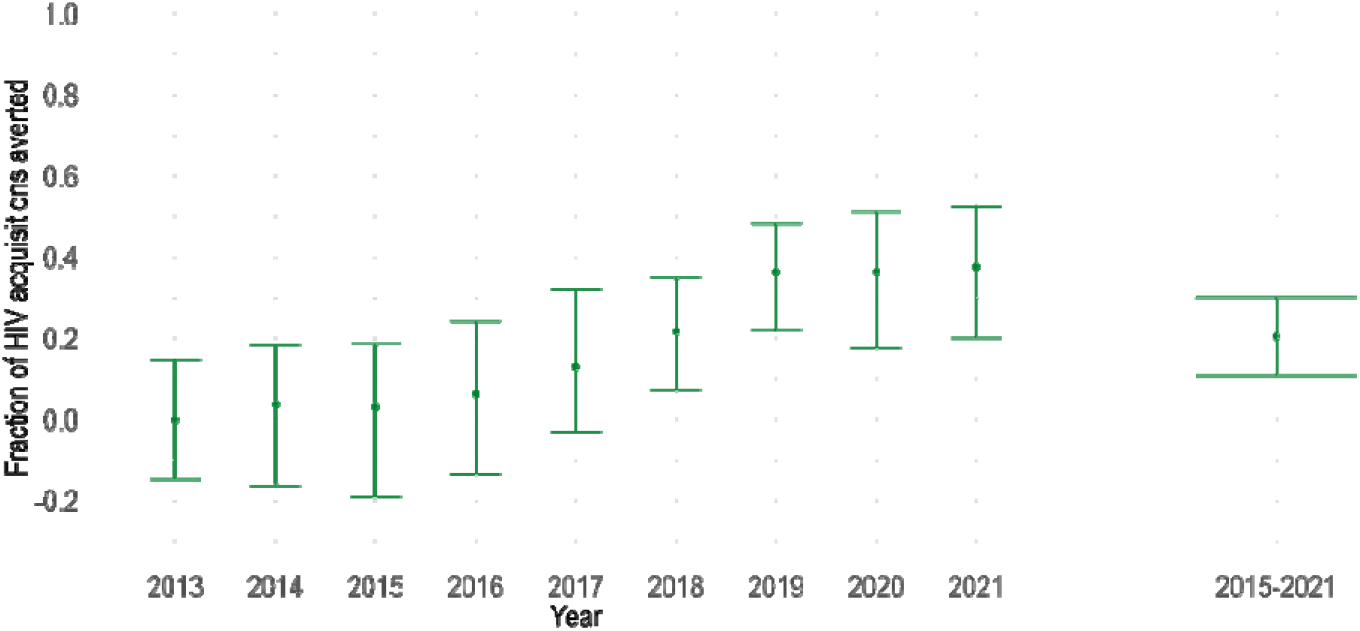
HIV Acquisitions averted under the provincial pre-exposure prophylaxis (PrEP) intervention scenario. Estimated annual (2013-2021) and cumulative (2015-2021) fractions of acquisitions averted due to the provincial PrEP intervention among men who have sex with men in Montréal. The coloured points and bars show the posterior mean and 90% credible intervals, respectively.

### Sensitivity Analyses

Our estimation of HIV incidence under PrEP intervention was not sensitive to the level of PrEP efficacy (86% vs 96%) or loosened eligibility criteria (Figure S7).

### Alternative PrEP intervention scenarios

Our alternative (hypothetical) analyses (Table 2) suggested that to have improved the impact of PrEP compared to the provincial intervention scenario, prioritizing MSM in the high sexual activity group (same overall coverage) or attaining higher overall PrEP coverage of 5% or 10% and 20% or 30% coverage by 2015 and 2019, respectively, would have been needed (Figure 5). Prioritizing PrEP to MSM in the high sexual activity group (same coverage as the provincial intervention) cumulatively averted 30% (90%CrI: 19%-42%) of HIV acquisitions over 2015-2021, with a peak annual fraction averted of 52% (90%CrI: 30%-69%) in 2021 (Figure 5). Even higher impacts could have resulted by reaching coverage targets of 5% or 10% by 2015 and 20% or 30% by 2019, especially when combined with prioritized use for MSM with high sexual activity levels. For instance, the scenario with the smallest increase in PrEP coverage, reaching targets of 5% in 2015 and 20% in 2019, averted 49% (90%CrI: 39%-57%) of HIV acquisitions between 2015-2022 (under provincial eligibility criteria), and up to 68% (90%CrI: 54%-70%) when prioritizing PrEP to the high sexual activity group. Conversely, the scenario with the largest increase in PrEP coverage, mirroring the coverage in Vancouver, reached targets of 10% by 2015 and 30% by 2019 and, under provincial eligibility criteria, averted 63% (90%CrI: 54%-70%) of HIV acquisitions over 2015-2021.

**Figure 5.**
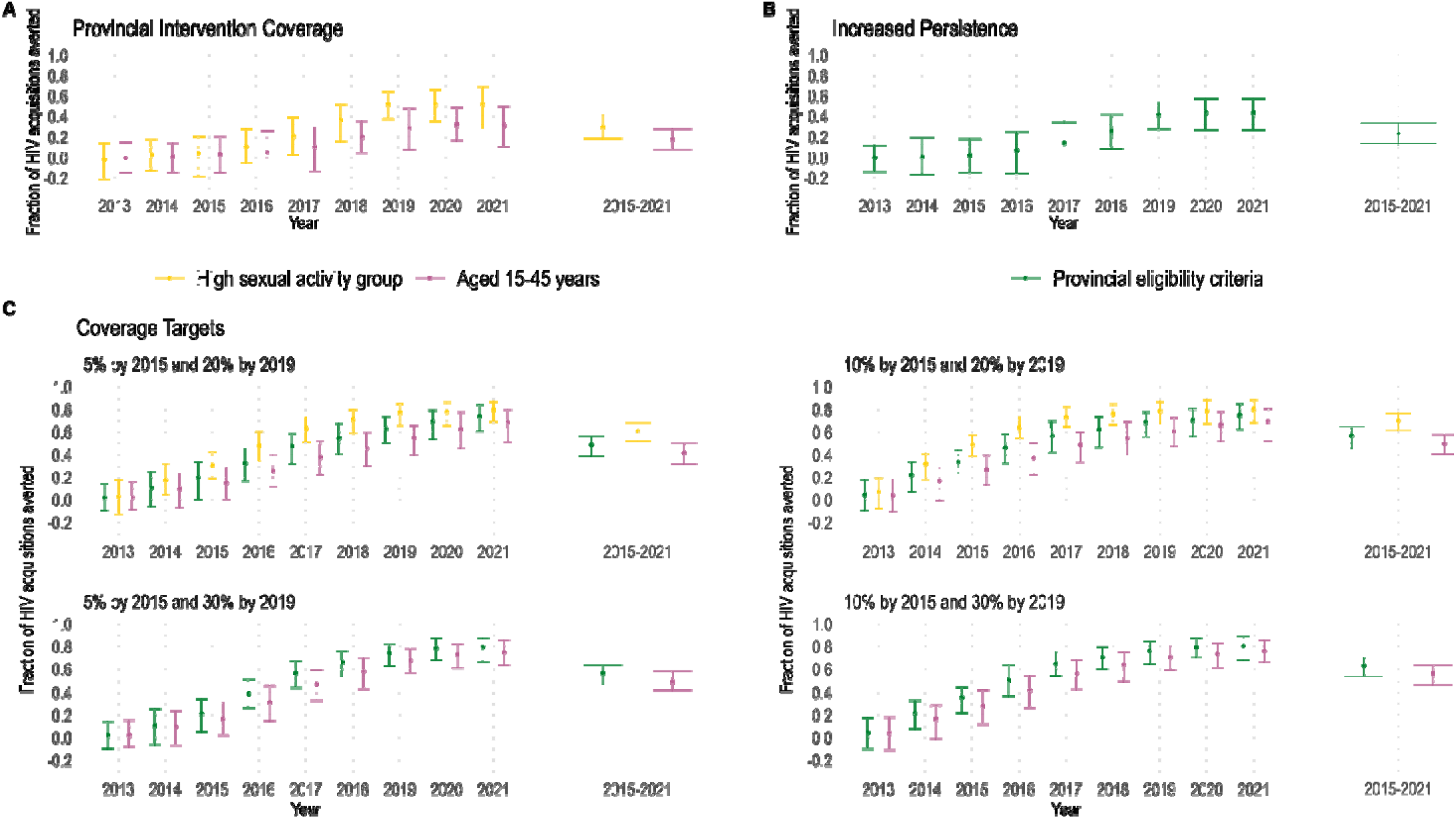
HIV Acquisitions Averted in Alternative Intervention Scenarios. Estimated fraction of acquisitions averted due to pre-exposure prophylaxis (PrEP) intervention among men who have sex with men (MSM) in Montréal under alternative (hypothetical) intervention scenarios. For each, the annual estimates from 2013-2021 and cumulative estimate over 2015-2021 are shown. The coloured points and bars show the posterior mean and 95% credible intervals, respectively. The panels display the results of: A) maintaining the observed PrEP coverage but prioritizing uptake in 1) the high sexual activity group or 2) those aged 15-45 years; B) maintaining the calibrated PrEP uptake probabilities but increasing retention on PrEP (for simplicity, only the results of the 50% decrease in discontinuation probability scenario are plotted); and C) increasing coverage up to a maximum of 30% by 2019 for three different uptake assumptions: 1) the same as the provincial PrEP eligibility criteria, or prioritizing uptake in 2) the high sexual activity group or 3) those aged 15-45 years. Note in the scenarios prioritizing MSM in the high sexual activity group it was not possible to reach 30% coverage by 2019 due to the insufficient number of individuals in the group. The results for these scenarios are not presented.

## Interpretation

This study presents a population-level estimate of PrEP’s impact, considering both direct and indirect effects. Using a detailed agent-based model of sexual HIV transmission and prevention among Montréal’s MSM, we created a valid counterfactual scenario without PrEP and found that, despite relatively low coverage, PrEP may have averted 20% (90%CrI: 11%-30%) of new HIV acquisitions between 2015-2022 in this population. From 2015-2019, as time and coverage accrued to 10% of MSM not living with HIV and 15% of the PrEP-eligible, the annual fraction of acquisitions averted by PrEP rose from 3% to 36%. Afterward, despite the COVID-19 pandemic disruptions to coverage, this level of impact persisted due to the transmission chains already prevented by PrEP.

Although our analysis suggests considerable population-level benefits of PrEP intervention among MSM in Montréal, it also highlights missed prevention opportunities. One obvious way to improve PrEP’s impact is to attain higher coverage. At the observed levels, prioritizing MSM with higher sexual activity levels might have improved impact, but not to the same extent as achieving higher coverage with the current provincial eligibility criteria, which approximately 60% of MSM not living with HIV met in our model. Increasing coverage to 5% or 10% in 2015 and 20% or 30% in 2019 could have prevented more than twice the number of HIV acquisitions since 2015. Given the coverage estimates over 2017-2020 for Vancouver MSM, we believe that levels of 10% in 2015 and 30% in 2019 could have been feasible in Montréal and would have averted an estimated 63% (90%CrI: 54%-70%) of HIV acquisitions would have been averted instead of 20%. Vancouver’s higher coverage has been attributed to total public funding for PrEP in that province^34^, whereas the PrEP co-payment in Québec can be as high as CAD$96.74 per month^18^. Even matching 5% coverage in 2015 and 20% in 2019 could have averted 49% (90%CrI: 39%-57%) of HIV acquisitions between 2015-2022, rising to 68% (90%CrI: 54%-70%) if efforts focused on MSM frequently engaging in anal sex with different partners.

Two other studies have examined the impact of PrEP intervention empirically using surveillance data and observed changes HIV diagnoses pre- and post-PrEP implementation^11, 12^. In Australia’s state of New South Wales, PrEP reached an estimated 20% of MSM living without HIV in 2016 and reduced new diagnoses of recent HIV acquisitions among MSM by 31.5% (95%CI: 11.3%-47.3%) in a 12-month post-implementation period, compared to the year prior^11^. A similar study in Scotland, where PrEP is free-of-cost from national clinics, estimated that 20% of MSM attending such clinics were prescribed PrEP and showed a 35.6% (95%CI: 7.1%-55.4%) reduction in new diagnoses of recent HIV acquisitions among MSM over a 24-month post-implementation period^12^.

It is challenging to directly compare these findings to ours due to different time frames (PrEP’s impact is expected to accrue over time) and variations in coverage and use. The most comparable period of our study is over 2017 and 2018 when coverage was beginning to rise. In those years, we estimated that 13% (90%CrI: 0%-32%) and 22% (90%CrI: 7%-35%) of acquisitions were averted, respectively. While differences between our estimates may stem from lower coverage in our study, heterogeneity in risk across the populations could also differ. Additionally, other factors like changes in HIV testing trends, the use of diagnoses of recent acquisitions to proxy incidence, and concomitant improvements in the treatment and care cascade could influence the findings of these studies.

Our analysis helped highlight missed opportunities in PrEP delivery in the city. For example, it took approximately two years for PrEP coverage to start increasing after Québec issued guidelines. Better awareness of PrEP’s efficacy and safety among healthcare providers and potential users following the early cessation of the IPERGAY trial in 2014 could have encouraged earlier use. Even after PrEP expanded, coverage maximized at 10%, never reaching the Vancouver’s levels. Our model indicated that many eligible MSM were not engaged in PrEP care yet could have benefited from its use, as evidenced by their higher HIV incidence. The financial burden of PrEP in Montréal remains an important barrier, potentially leaving behind those most at risk of HIV acquisition^35^.

The findings from our impact evaluation should be interpreted cautiously. Our model used simplifying assumptions about disease progression, HIV transmission, and prevention use. For instance, individuals on PrEP may be less likely to serosort (i.e., choose partners not living with HIV), and PLHIV may preferentially mix with PrEP users^30, 36^. If mixing was less assortative by HIV status among those on PrEP, we might have slightly overestimated the impact of PrEP. However, this potential bias should be small given the high viral suppression levels. Additionally, the model did not consider risk compensation or changes in sexual behavior that could be associated with PrEP use, which could overestimate the impact of PrEP. However, this overestimation should be small, given PrEP’s effect size. Finally, the model did not differentiate between daily and on-demand regimens, given their equivalent efficacy under perfect adherence.

Our analysis has several strengths. Firstly, we leveraged data from multiple cohorts and surveys of MSM in Montréal to parameterize and calibrate the model. Secondly, using a model allowed us to construct an appropriate counterfactual scenario without a suitable control group^13^. Thirdly, we controlled for changes in Québec’s treatment eligibility criteria. Lastly, our impact estimates include PrEP’s direct effects in preventing HIV acquisition among users and the indirect benefits to MSM not on PrEP due to overall decreases in HIV prevalence.

## Conclusion

PrEP has the potential to contribute significantly to HIV elimination as a key part of combination HIV prevention. However, global scale-up has been slow, resulting in sub-optimal coverage and limited impact on incidence reductions^37^. In Montréal, despite modest coverage, PrEP had a notable impact on HIV transmission, complementing declining incidence and high ART coverage. Free PrEP could remove important barriers, but more needs to be done to address stigma, discrimination, and certain physicians’ reticence to prescribe PrEP, amongst others^38-41^. The availability of alternative formulations like long-acting injectable PrEP could further usage and support adherence. By removing barriers, we can accelerate HIV elimination among MSM and other vulnerable populations.

## Data Availability

The data that support the findings of this study are available from Engage‐ Montréal but restrictions apply to the availability of these data, which were used under license for the current study, and so are not publicly available. Data are however available from the authors upon reasonable request and with permission of Engage‐Montréal.

## Notes

**Financial sponsors:** CMD is supported by a doctoral award from the *Fonds de recherche du Québec – Santé* (FRQS). Grants from the *Canadian Foundation for AIDS Research* and the *Canadian Institutes of Health Research* (CIHR) to MM-G. MM-G’s research program is funded by the *Tier 2 Canada Research Chair in Population Health Modelling*. SM is supported the *Tier 2 Canada Research Chair in Mathematical Modelling and Program Science* and the *Ontario HIV Treatment Network*. CT is the *Pfizer/Université de Montréal Chair in HIV Translational Research*. MCB acknowledges funding from the *MRC Centre for Global Infectious Disease Analysis* (reference MR/R015600/1), jointly funded by the *UK Medical Research Council* (MRC) and the *UK Foreign, Commonwealth & Development Office* (FCDO), under the MRC/FCDO Concordat agreement and is also part of the EDCTP2 programme supported by the European Union. For the purpose of open access, MCB has applied a Creative Commons Attribution (CC BY) license to any Author Accepted Manuscript version arising. DG is supported by the *Tier 2 Canada Research Chair in Sexual and Gender Minority Health Research*.

**Conflict of interest statement:** JC has investigator-sponsored research grants from Gilead Sciences Canada and ViiV Healthcare. He has also received financial support for conference travel and advisory work for Gilead Sciences Canada, Merck Canada and ViiV Healthcare. MM-G reports an investigator-sponsored research grant from *Gilead Sciences Inc*., contractual arrangements from the *World Health Organization*, the *Joint United Nations Programme on HIV/AIDS* (UNAIDS), the *Institut national de santé publique du Québec* (INSPQ), and *the Institut d’excellence en santé et services sociaux* (INESSS), all outside of the submitted work. CT has investigator-sponsored research grants from Merck and Gilead Sciences Canada, and has received financial support for advisory work and conferences from Gilead Sciences Canada, Merck, Medicago, Astra-Zeneca, Pfizer, Sanofi, and GSK. J-GB has received honoraria for consulting for ViiV, Healthcare, Merck, and Gilead Sciences Canada and for participation as a speaker at conferences supported by Merck and Gilead Sciences unrelated to this work.

### Competing Interest Statement

JC has investigator-sponsored research grants from Gilead Sciences Canada and ViiV Healthcare. He has also received financial support for conference travel and advisory work for Gilead Sciences Canada, Merck Canada and ViiV Healthcare. MM-G reports an investigator-sponsored research grant from Gilead Sciences Inc., contractual arrangements from the World Health Organization, the Joint United Nations Programme on HIV/AIDS (UNAIDS), the Institut national de santé publique du Québec (INSPQ), and the Institut d'excellence en santé et services sociaux (INESSS), all outside of the submitted work. CT has investigator-sponsored research grants from Merck and Gilead Sciences Canada, and has received financial support for advisory work and conferences from Gilead Sciences Canada, Merck, Medicago, Astra-Zeneca, Pfizer, Sanofi, and GSK. J-GB has received honoraria for consulting for ViiV, Healthcare, Merck, and Gilead Sciences Canada and for participation as a speaker at conferences supported by Merck and Gilead Sciences unrelated to this work. RéZO (AD-B) has received sponsorships from Gilead Sciences Canada.

### Funding Statement

CMD is supported by a doctoral award from the Fonds de recherche du Québec-Santé (FRQS). Grants from the Canadian Foundation for AIDS Research and the Canadian Institutes of Health Research (CIHR) to MM-G. MM-G's research program is funded by the Tier 2 Canada Research Chair in Population Health Modelling. SM is supported the Tier 2 Canada Research Chair in Mathematical Modelling and Program Science and the Ontario HIV Treatment Network. CT is the Pfizer/Université de Montréal Chair in HIV Translational Research. MCB acknowledges funding from the MRC Centre for Global Infectious Disease Analysis (reference MR/R015600/1), jointly funded by the UK Medical Research Council (MRC) and the UK Foreign, Commonwealth & Development Office (FCDO), under the MRC/FCDO Concordat agreement and is also part of the EDCTP2 programme supported by the European Union. For the purpose of open access, MCB has applied a Creative Commons Attribution (CC BY) license to any Author Accepted Manuscript version arising. DG is supported by the Tier 2 Canada Research Chair in Sexual and Gender Minority Health Research.

### Author Declarations

The IRB of the McGill University Research Ethics Board gave ethical approval for this work. The IRB of the Research Institute of the McGill University Health Centre gave ethical approval for this work.

