## Supplementary Materials for "Population-level effectiveness of pre-exposure prophylaxis for HIV prevention among men who have sex with men in Montréal: a modelling study of surveillance and survey data"

Carla M Doyle^1^, Rachael M Milwid^1^, Joseph Cox^1,2,3^, Yiqing Xia^1^, Gilles Lambert^2^, Cécile Tremblay^4,5^, Joanne Otis^6^, Marie-Claude Boily^7^, Jean-Guy Baril^8,9^, Sarah-Amélie Mercure^2^, Réjean Thomas^10^, Benoit Trottier^9^, Sharmistha Mishra^11, 12, 13^, Mathieu Maheu-Giroux^1§^

^1^Department of Epidemiology and Biostatistics, School of Population and Global Health, McGill University, Montréal, QC

^2^Direction Régionale de Santé Publique de Montréal, Montréal, QC

^3^Clinical Outcomes Research and Evaluation, Research Institute - McGill University Health Centre, Montréal, QC

^4^Centre de Recherche du Centre Hospitalier de l'Université de Montréal (CRCHUM), Montréal, QC

^5^Département de Microbiologie, Infectiologie et Immunologie, Université de Montréal, Montréal, QC

^6^Département de Sexologie, Université du Québec à Montréal, Montréal, QC

^7^Department of Infectious Diseases, Imperial College London, London, UK

^8^Department of Family Medicine, Centre Hospitalier de l’Université de Montréal, Montréal, QC

^9^Clinique de médecine urbaine du Quartier Latin, Montréal, QC

^10^Clinique médicale l'Actuel, Montréal, QC

^11^Department of Medicine, St. Michael's Hospital, University of Toronto, Toronto, ON

^12^Institute of Medical Sciences, University of Toronto, Toronto, ON

^13^Institute of Health Policy Management and Evaluation, Dalla Lana School of Public Health, University of Toronto, Toronto, ON

### 1. Pre-exposure prophylaxis (PrEP) parameterization: Additional details

##### Eligibility

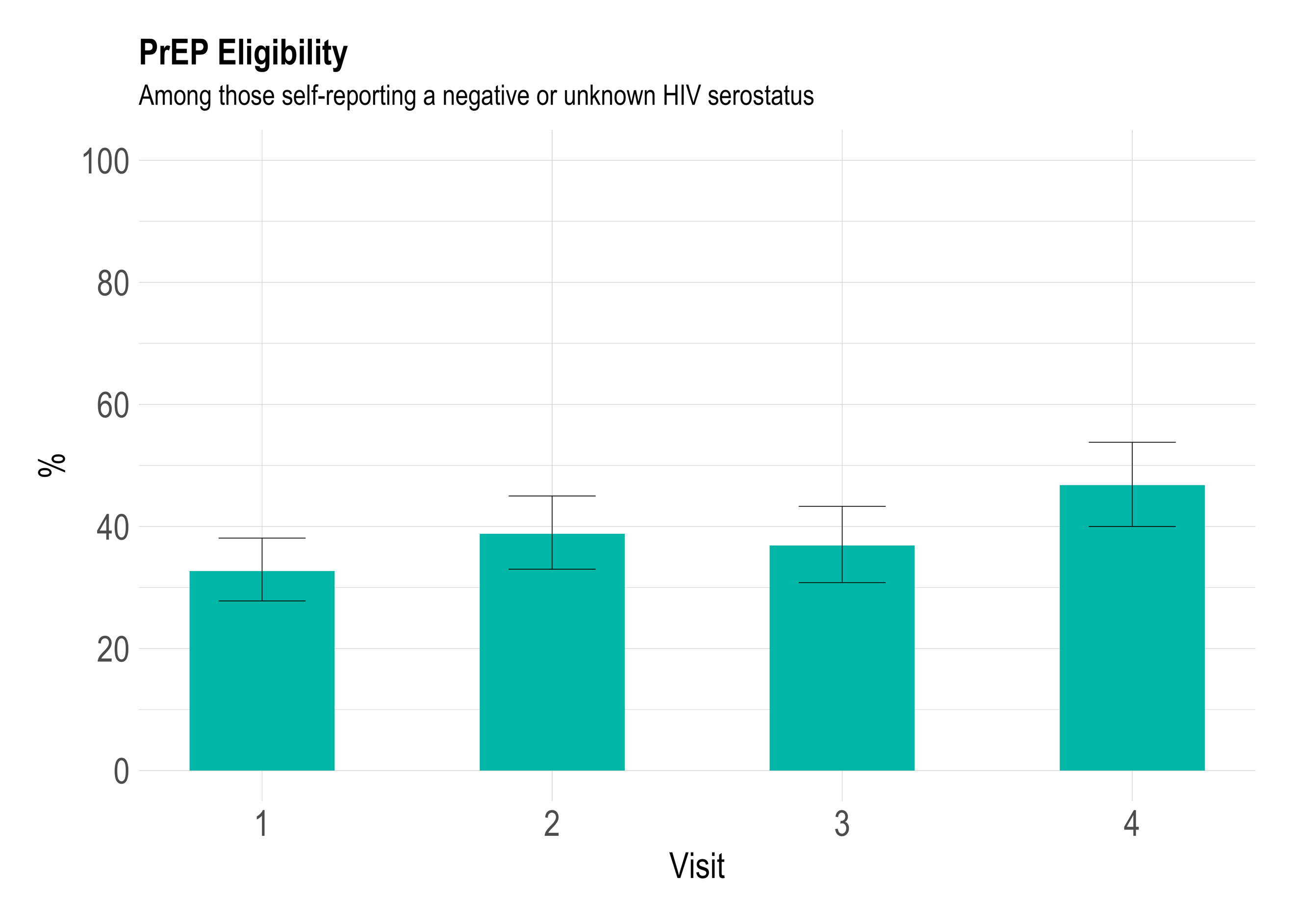

**Figure S1.** **Empirical estimates of pre-exposure prophylaxis (PrEP) eligibility among men who have sex with men (MSM) in Montréal.** The estimated annual percentage of Montréal MSM eligible for PrEP (according to the modelled provincial criteria) among Engage participants that self-reported a negative or unknown HIV serostatus. The first four Engage study visits occurred annually over 2017-2021. All estimates were adjusted by RDS-II and inverse probability of censoring weights. The error bars show the estimated 95% confidence intervals.

##### Initiation

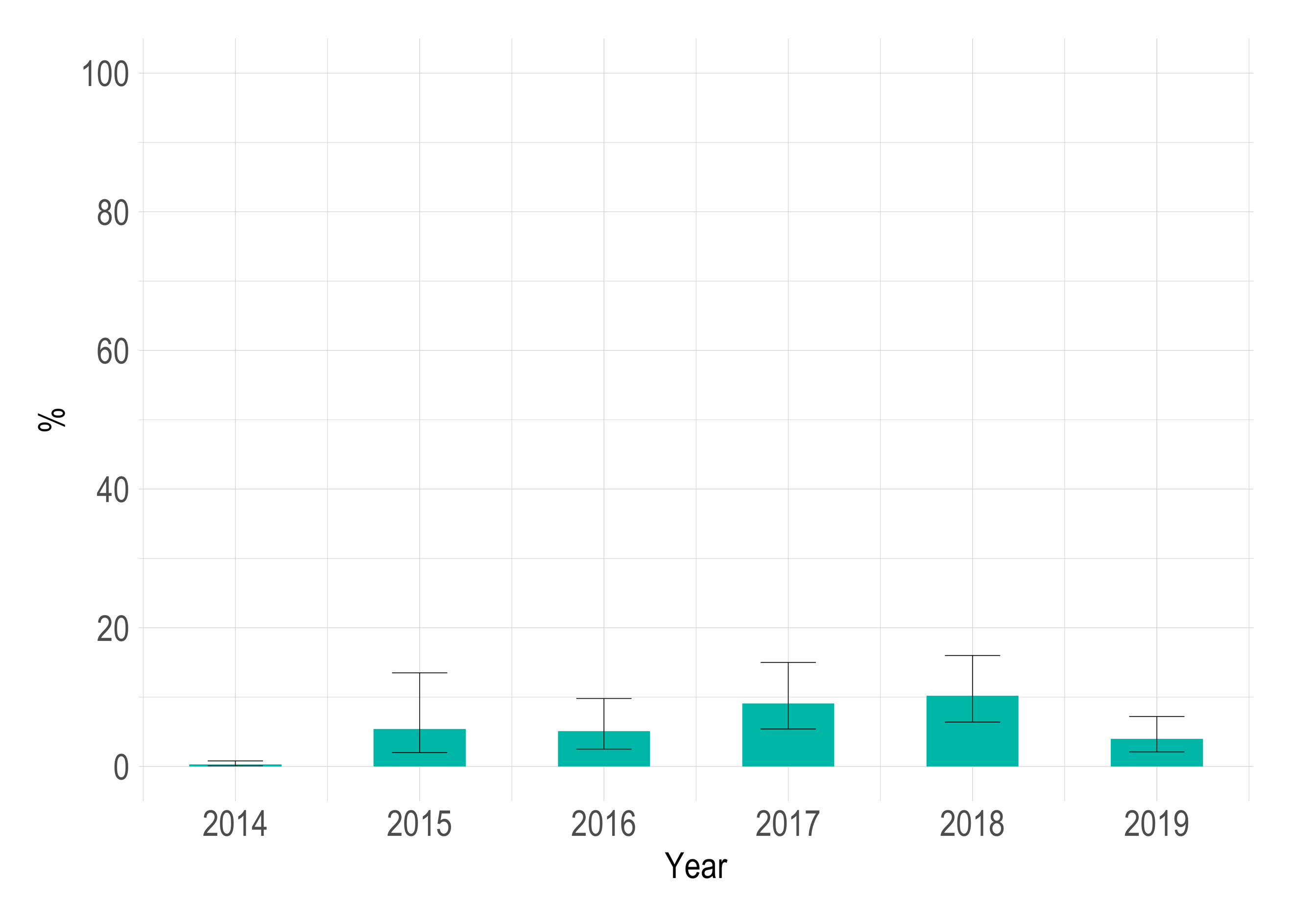

**Figure S2. Empirical estimates of pre-exposure prophylaxis (PrEP) uptake among men who have sex with men (MSM) in Montréal.** The estimated percentage of Montréal MSM that reported first taking PrEP in a given year among Engage participants eligible for PrEP (according to the modelled provincial criteria). No Engage participant reported first taking PrEP in 2013. To obtain estimates before 2017 (when the Engage cohort study began), we assumed the number eligible at baseline was constant. All estimates were adjusted by RDS-II and inverse probability of censoring weights. The error bars show the estimated 95% confidence intervals.

##### Adherence

*Engage* measured adherence among continuous PrEP users (those reporting ever using PrEP continuously at baseline or in the past six months throughout follow-up) at all visits by capturing the self-reported number of daily doses missed per week (*“On average, how many days per week have you missed your dose of PrEP medication?”*). Additionally, a measure of PrEP-protected anal sex was included in the study questionnaire as of the third visit (*“In the past 6 months, how often were you on PrEP when you had sexual activities involving anal sex (either as top or bottom)?”*). Together, these measures indicated consistently high levels of self-reported adherence among MSM taking PrEP daily (Figure S3). However, across visits, approximately 40%-60% of Engage participants reporting PrEP use self-reported strictly continuous use (Figure S4). Therefore, many Montréal MSM do indeed use PrEP on a situational basis. Without understanding the adherence or pill taking frequency of on-demand users in our setting, we did not model differential PrEP adherence. Instead, we parameterized PrEP effectiveness by the intention-to-treat estimate from the IPERGAY trial (which included Montréal MSM)^1^. Among all IPERGAY participants, 43% (95%CI: 35%-51%) self-reported correct use of the on-demand PrEP schedule^1^. In sensitivity analyses, we modelled an increased efficacy of 96%, corresponding to taking four doses per week, the threshold often used to define high adherence^2, 3^.

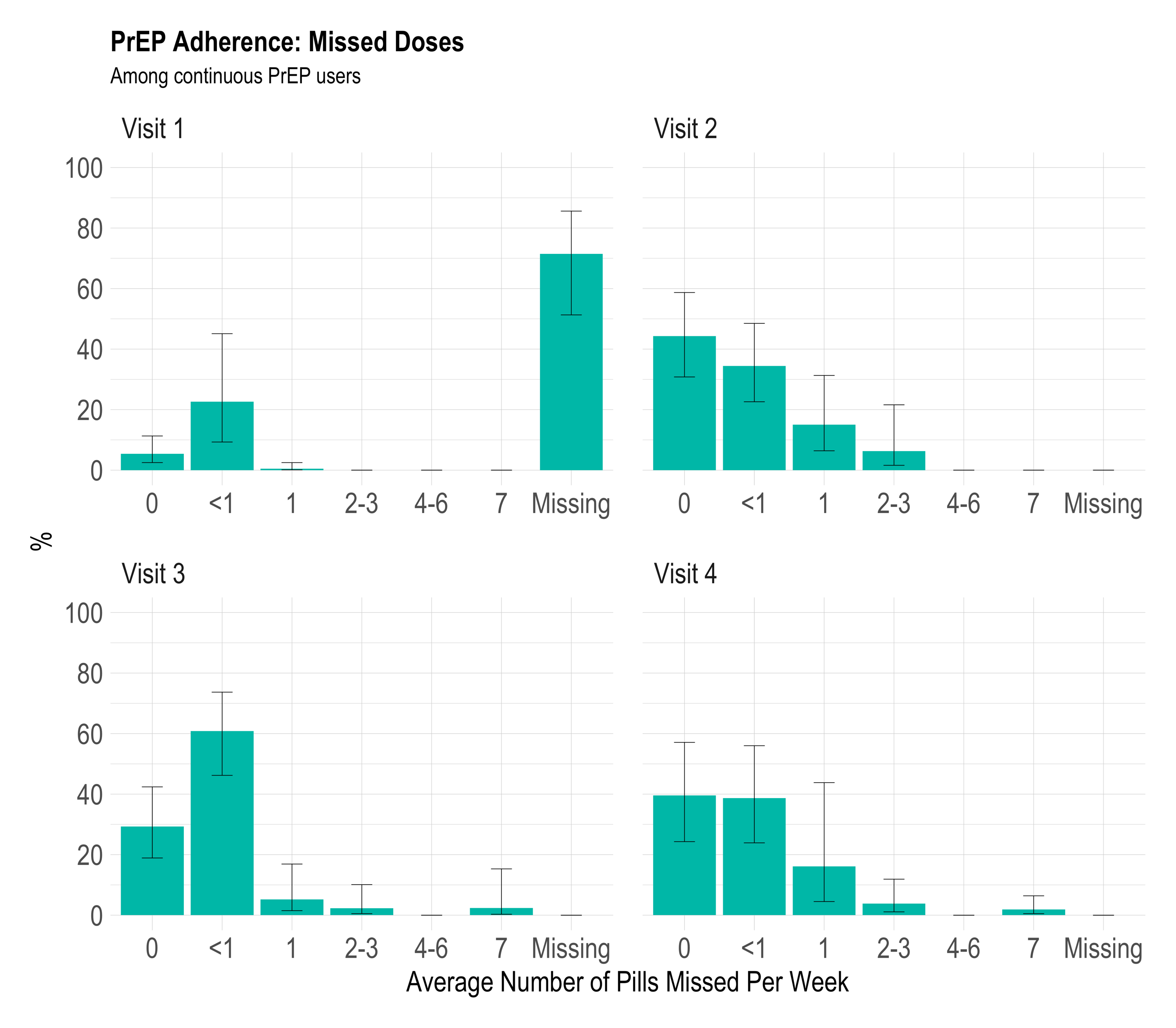
**A**

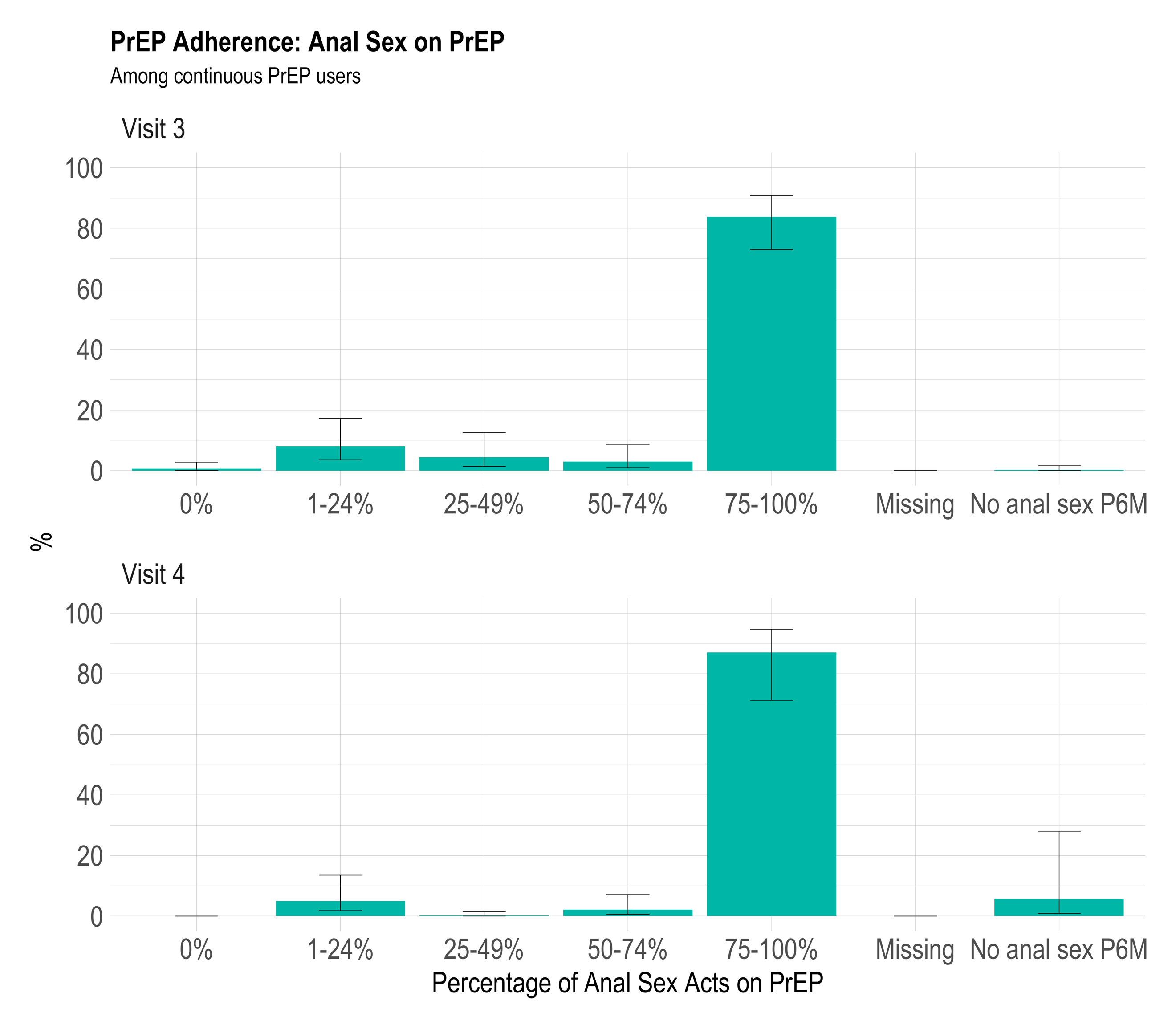
**B**

**Figure S3.** **Empirical estimates of pre-exposure prophylaxis (PrEP) adherence among men who have sex with men (MSM) in Montréal.** Estimates of PrEP adherence among Montréal MSM calculated using the Engage cohort and adjusted by RDS-II and inverse probability of censoring weights. The first four Engage study visits occurred annually over 2017-2021. Panel A displays the self-reported average number of pills missed per week among continuous PrEP users at each study visit. Panel B displays the self-reported percentage of anal sex acts covered by PrEP among continuous PrEP users at the third and fourth study visits. The error bars show the estimated 95% confidence intervals.

##### Schedule

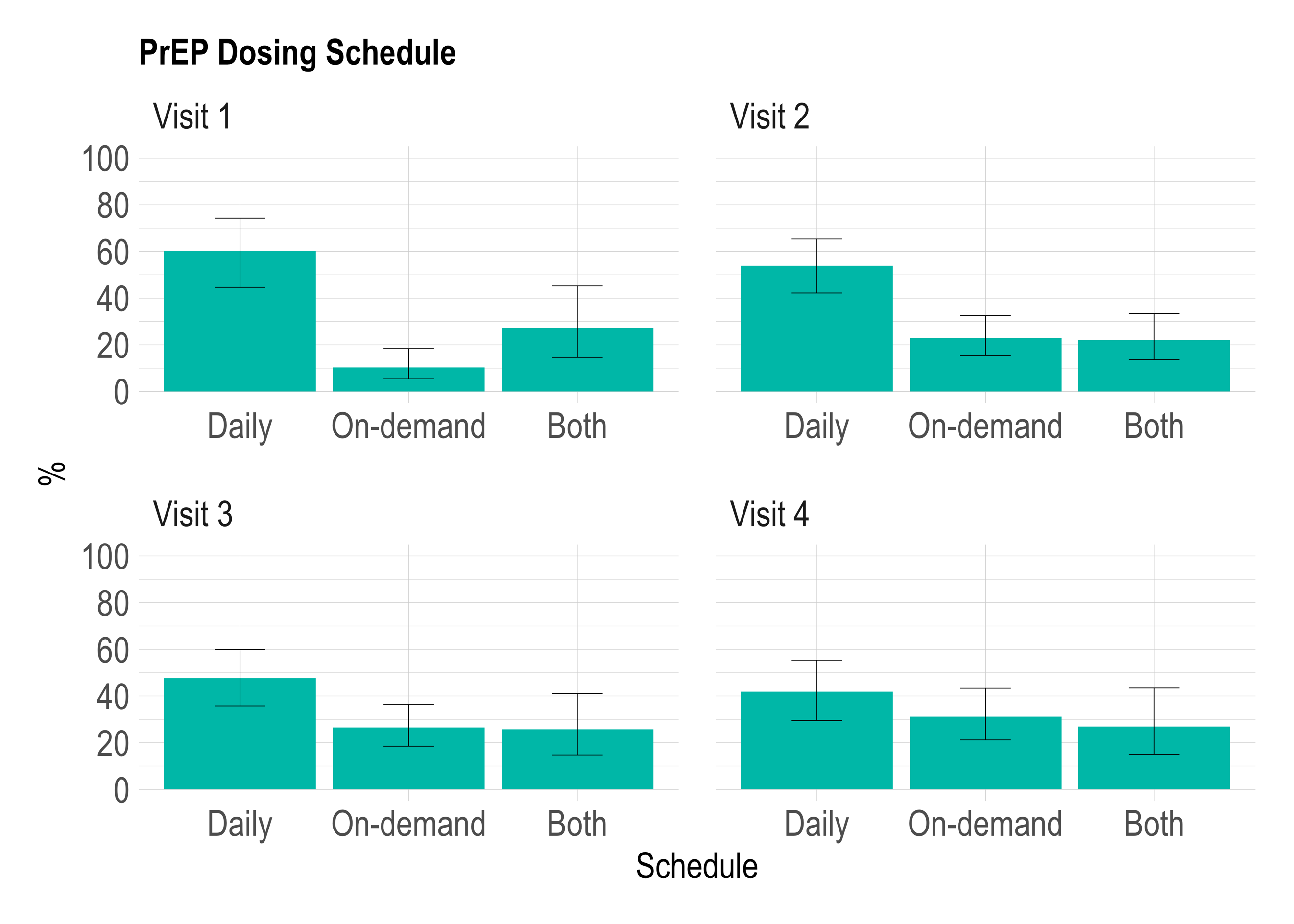

**Figure S4.** **Empirical estimates of pre-exposure prophylaxis (PrEP) dosing schedule among men who have sex with men (MSM) in Montréal.** The estimated percentage of PrEP users following a daily or on-demand dose schedule calculated using the Engage cohort and adjusted by RDS-II and inverse probability of censoring weights. The error bars show the estimated 95% confidence intervals. The first four Engage study visits occurred annually over 2017-2021.

### 2. Additional Model Results

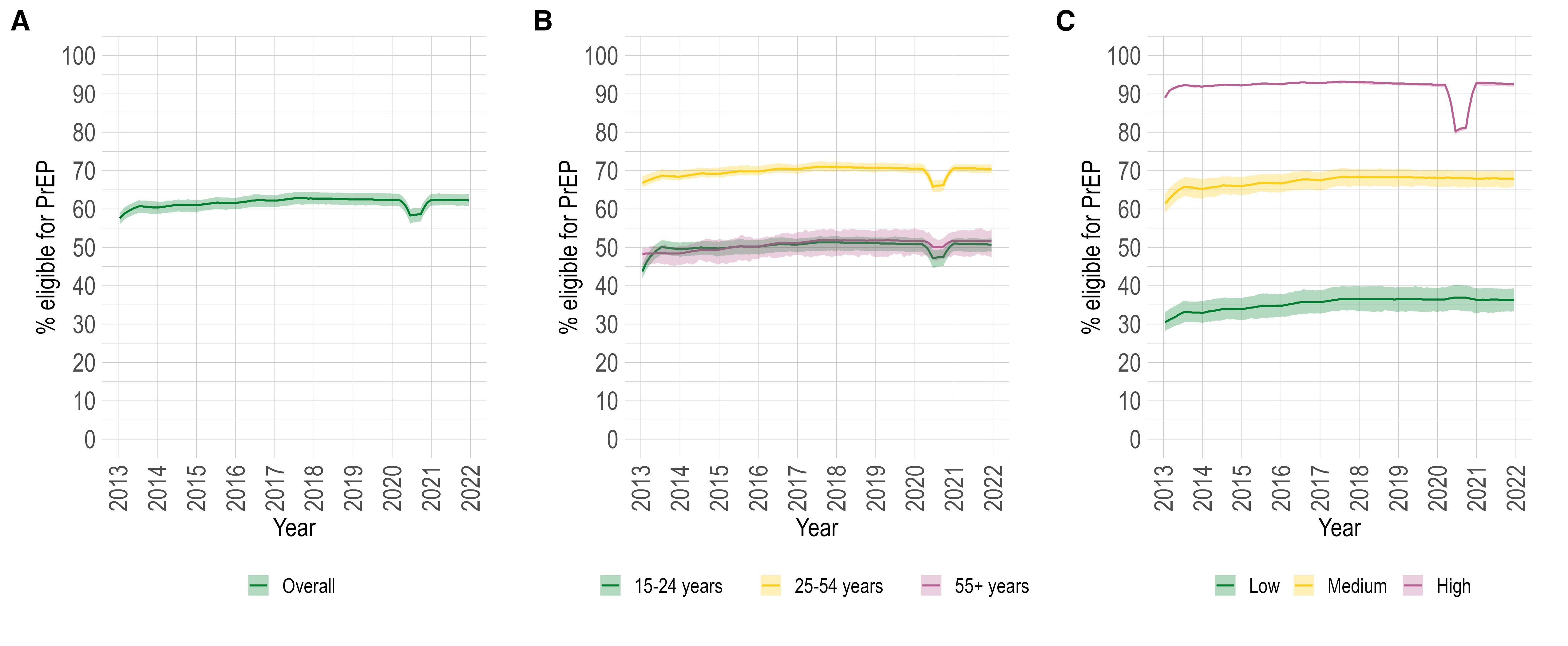

**Figure S5. Modelled pre-exposure prophylaxis (PrEP) eligibility among men who have sex with men (MSM) not living with HIV in Montréal.** The model estimated percentage of MSM not living with HIV eligible for PrEP over 2013-2021 in Montréal: overall (panel A) and stratified by age (panel B) and sexual activity group (panel C). The coloured lines and bands show the model posterior mean and 90% credible intervals, respectively.

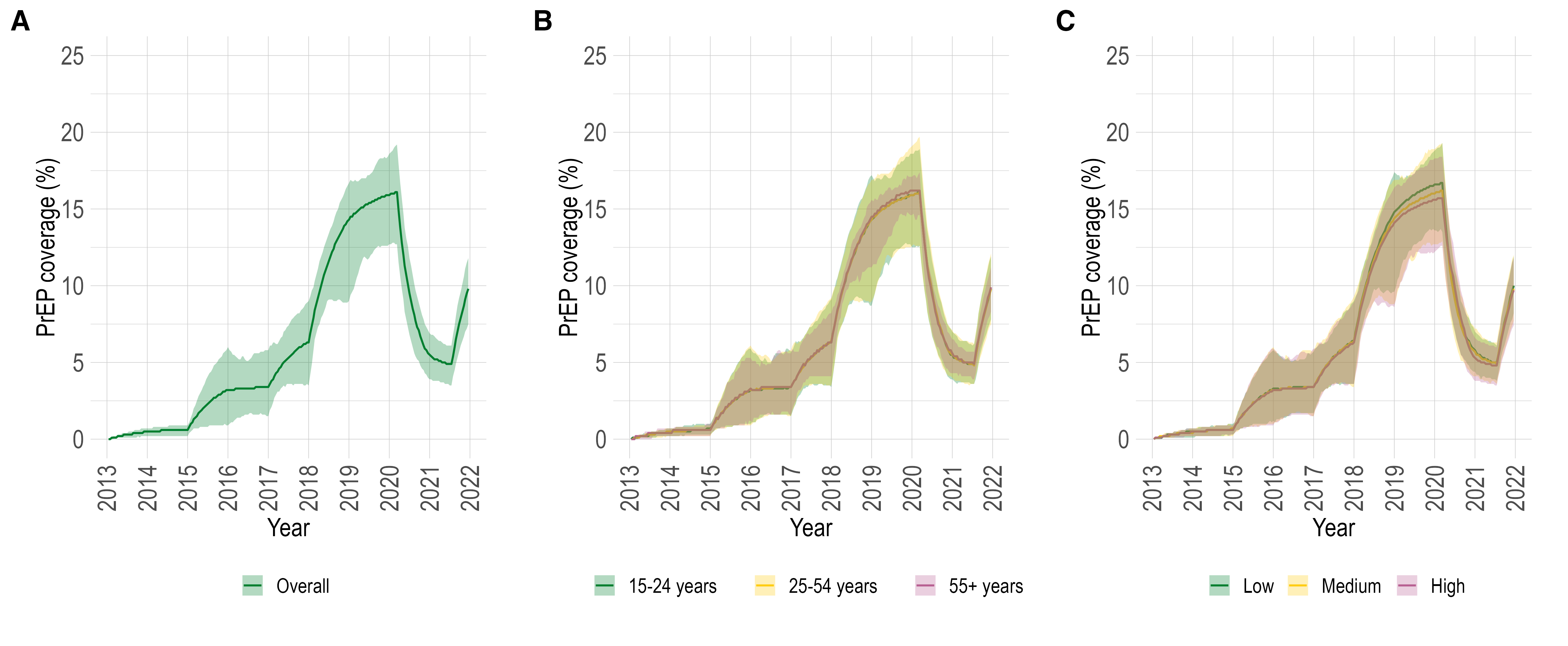

**Figure S6. Modelled pre-exposure prophylaxis (PrEP) coverage among PrEP-eligible men who have sex with men (MSM) not living with HIV in Montréal.** The model estimated PrEP coverage over 2013-2021 among MSM eligible for PrEP in Montréal: overall (panel A) and stratified by age (panel B) and sexual activity group (panel C). The coloured lines and bands show the model posterior mean and 90% credible intervals, respectively.

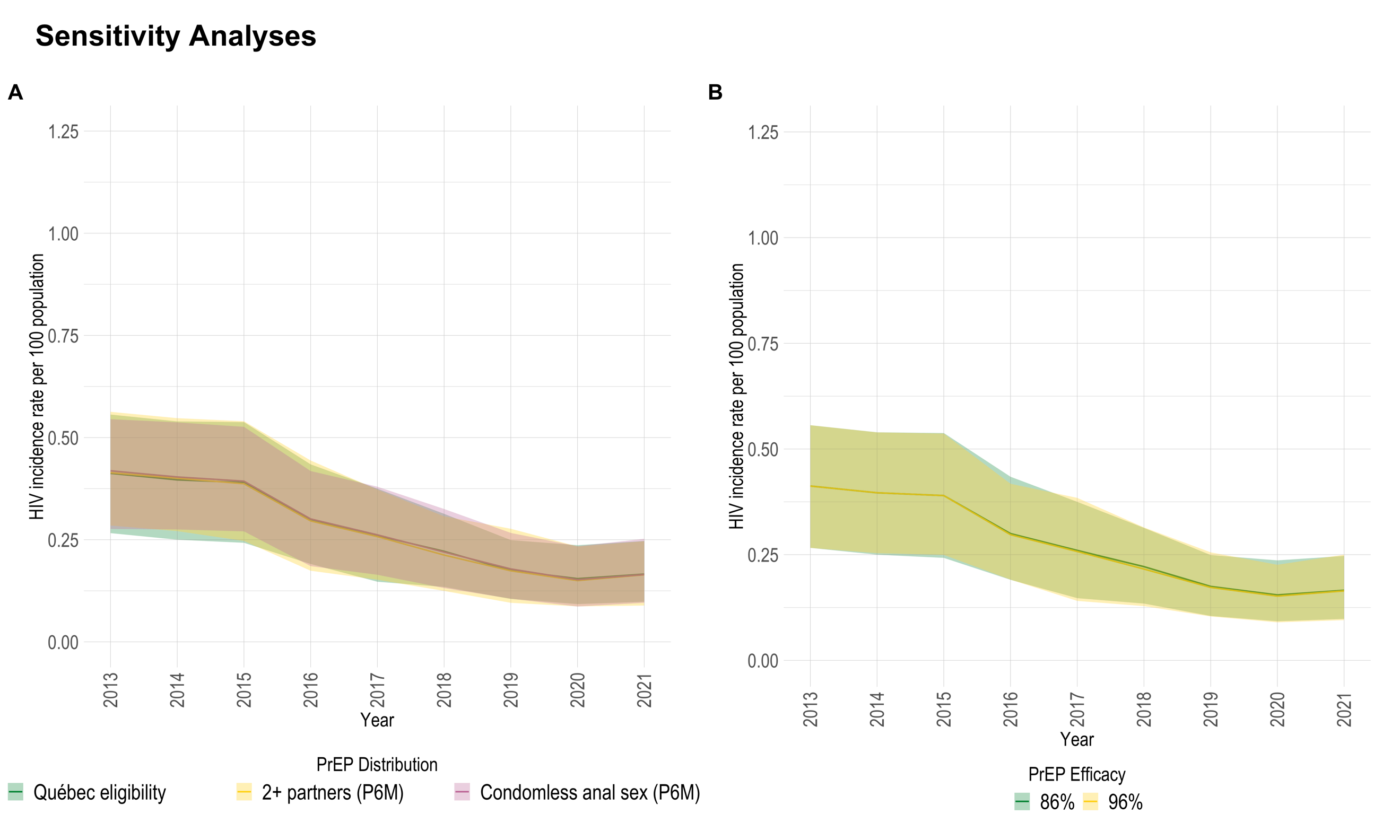

**Figure S7. Sensitivity Analyses.** The model estimated HIV incidence rates over 2013-2021 among Montréal men who have sex with men (MSM) under the provincial pre-exposure prophylaxis (PrEP) intervention scenario with different PrEP-eligibility criteria (panel A) and with different PrEP efficacies (Panel B). The coloured lines and bands show the posterior median and 90% credible intervals, respectively.

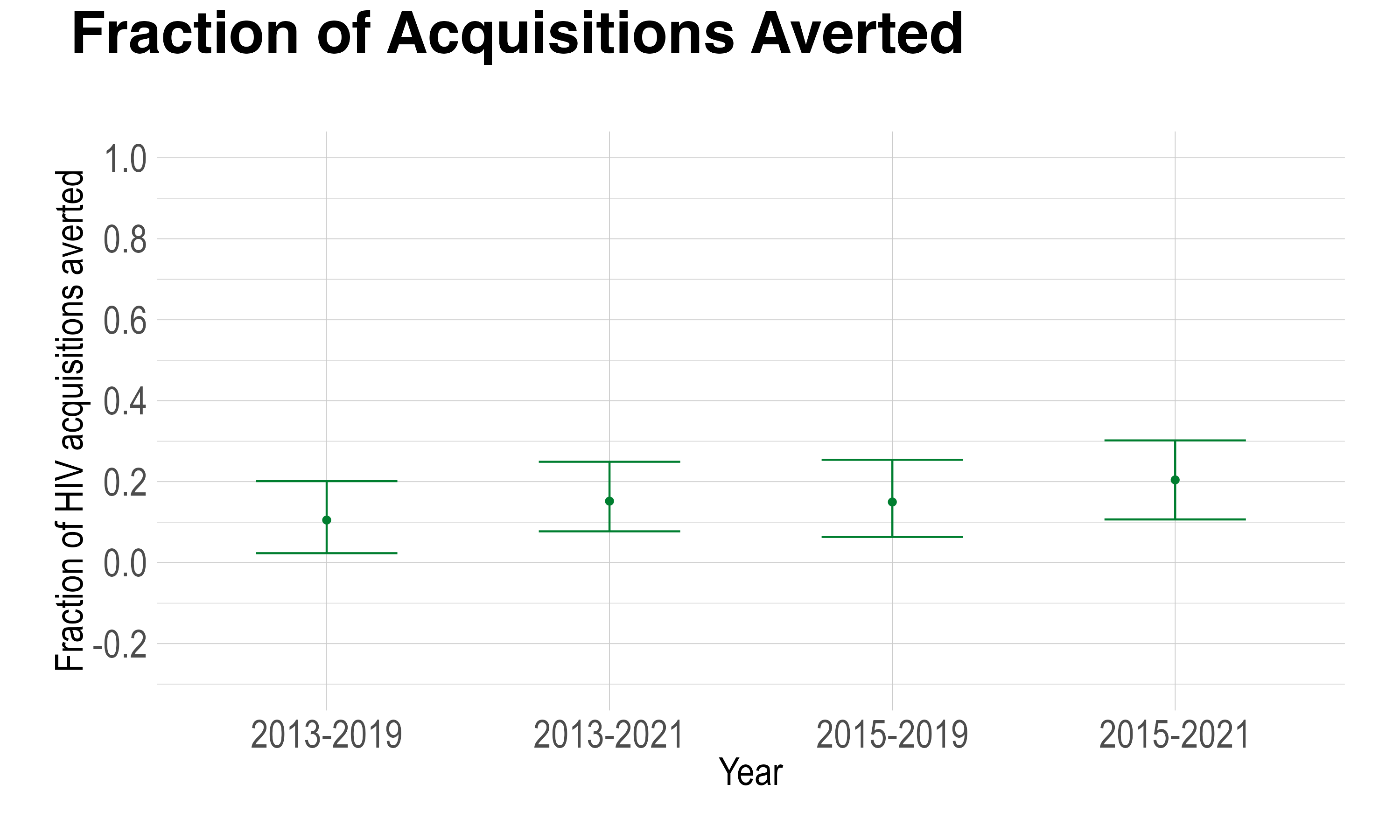

**Figure S8. Cumulative Acquisitions Averted.** Estimated cumulative fraction of acquisitions averted due to pre-exposure prophylaxis (PrEP) intervention among men who have sex with men (MSM) in Montréal (provincial PrEP intervention scenario) over varying time periods. The coloured points and bars show the posterior mean and 90% credible intervals, respectively.
